# Plasma glycosaminoglycans and cell-free DNA to discriminate benign and malignant lung diseases

**DOI:** 10.1101/2024.07.01.24309751

**Authors:** Alvida Qvick, Sinisa Bratulic, Jessica Carlsson, Bianca Stenmark, Christina Karlsson, Jens Nielsen, Francesco Gatto, Gisela Helenius

**Author notes:** Correspondence to: Alvida Qvick Adress: Dep. of Laboratory Medicine, Örebro University Hospital, Södra Grev. Roseng., 701 85 Örebro, Sweden. All authors have read and approved the final version of this manuscript. **Conflict of interest:** F. Gatto and J. Nielsen are shareholders in Elypta AB. F. Gatto and S. Bratulic are employed at Elypta AB. J. Nielsen is a board member at Elypta AB. Elypta AB has a commercial interest in part of the technology described in this study. Remaining authors have no conflicts to declare.

## Abstract

We aimed to investigate the use of free glycosaminoglycan profiles (GAGomes) and cfDNA in plasma to differentiate between lung cancer and benign lung disease. GAGs were analyzed using the MIRAM® Free Glycosaminoglycan Kit with ultra-high-performance liquid chromatography and electrospray ionization triple-quadrupole mass spectrometry. We detected two GAGome features, 0S chondroitin sulfate (CS) and 4S CS, with cancer-specific changes. Based on the observed GAGome changes, we devised a model to predict lung cancer. The model, named the GAGome score, could detect lung cancer with 41.2% sensitivity (95% CI: 9.2-54.2%) at 96.4% specificity (CI: 95.2-100.0%, n=113). Furthermore, we found that the GAGome score, when combined with a cfDNA test, could increase the sensitivity for lung cancer from 42.6% (95% CI: 31.7-60.6%, cfDNA alone) to 70.5% (CI: 57.4 - 81.5%) at 95% specificity (CI: 75.1-100%, n=74). Notably, the combined GAGome and cfDNA testing improved the sensitivity, especially in early stages, relative to the cfDNA alone. Our findings show that plasma GAGome profiles can enhance cfDNA testing performance, highlighting the applicability of a multiomics approach in lung cancer diagnostics.

## 1. Introduction

The use of high-throughput sequencing to analyze the genome and its aberrations has advanced diagnostics, predictive testing, and monitoring for many types of cancer. To avoid the challenges of obtaining tissue biopsies for cancers like lung cancer (LC), a liquid biopsy has emerged as a promising alternative. Numerous articles have been published on sequencing of circulating cell-free DNA (cfDNA) for such cancers^1^. Despite technical advances, which improved the sensitivity of genomics-only liquid biopsy assays, many cancers remain undetected. This can be due to technical as well as biological reasons since the released amount of circulating tumor DNA is very heterogeneous between different tumors^2^. To overcome this limitation, the field has turned beyond genomics to a combination of different omics techniques, referred to as multiomics, including combined information from genomics, transcriptomics, proteomics, methylomics, metabolomics, extracellular vesicles, and circulating tumor cells among others^3–8^.

Glycosaminoglycans (GAGs) are unbranched linear polysaccharides that can be divided into four main classes: chondroitin sulfate, heparin sulfate, keratan sulfate, and hyaluronic acid^9^. The sulfation and glycosylation patterns in GAG chains can vary widely, leading to a very high structural and functional diversity of resulting GAG molecules^10^. GAGs play crucial roles in cellular functions, including maintaining the extracellular matrix structure and providing hydration to cells. They are also integral to the immune response^11,12^, tissue homeostasis, as well as cell growth, proliferation, differentiation, and adhesion^13^. Notably, GAGs have been implicated in different aspects of cancer development and progression due to their involvement in the tumor microenvironment, through their interaction with growth factors, growth factor receptors, and cytokines^14^. The disaccharide composition of GAGs in tumor tissue, plasma, and urine has been shown to be altered in several cancer types such as breast, prostate, gastric, and renal cell carcinoma^15–19^. Conversely, only a few studies measured the structural profile of GAG disaccharides (or GAGome) in tissue or liquid biopsies in LC^20,21^, indicating the need for further research.

This study aimed to explore the plasma GAGome’s potential for differentiating LC from non-malignant lung diseases via liquid biopsy. Additionally, we investigated if combining GAGome and cfDNA data could increase the sensitivity and specificity to detect LC.

## 2. Materials and Methods

### 2.1 Study design and cohort characteristics

This study was reported in compliance with the Standards for Reporting of Diagnostic Accuracy (STARD) guidelines^22^ (Table S1). Study participants were enrolled into the study during routine clinical investigation, at the lung clinic at Örebro University Hospital, between February 2016 and February 2019. Patients investigated for the suspicion of LC were included in the study and formed a consecutive series. To ensure the most accurate diagnosis, all LC cases were histologically confirmed on tumor tissue. Patients diagnosed with cancer of other origin than lung, sample not collected prior to treatment start, or with inadequate tumor material for diagnosis, were excluded from the study. Participants gave written informed consent before inclusion and the study was approved by the regional ethics committee board in Uppsala (Approval 2015-400, 2021-01478).

Tumors were staged and histologically classified according to the guidelines of the International Association for the Study of Lung Cancer (IASLC) and the World Health Organization nomenclature, respectively^23,24^. Patients with benign lung diseases mainly consisted of different pulmonary obstructive diseases, inflammatory conditions in the lung, fibrosis in the lung or benign lung nodules.

### 2.2 Plasma collection and isolation

Blood was collected in Cell-Free RNA^TM^ BCT tubes (Streck, Omaha, NE) and plasma was retrieved by a two-step centrifugation; 2000×g for 10 min followed by 16 000×g for 10 min. Plasma was stored at -80° C until preparation for analysis. Samples were thawed on ice and an aliquot of each sample was taken for further analyses.

### 2.3 GAGome measurements

Plasma GAGome measurements were performed retrospectively in a single blinded Good Laboratory Practice (GLP)-compliant central laboratory using MIRAM® Free Glycosaminoglycan Kit (Elypta AB, Sweden), which is a standardized kit for GAG extraction, detection, and quantification by ultra-high-performance liquid chromatography (UHPLC) coupled with electrospray ionization triple-quadrupole mass spectrometry system (ESI-MS/MS, Acquity I-class Plus Xevo TQ-S micro, Waters® Corporation, MA). A single UHPLC column equipped with a pre-column guard (Waters® ACQUITY UPLC BEH C18 VanGuard Pre-column) was sufficient to analyze all samples in this study with no quality deterioration observed over time. The analytical performance characteristics of the kit have been previously described^25^.

In short, the kit is based on a method by Volpi *et al*. (2014)^26^. The assay consists of the enzymatic depolymerization of GAGs from the sample into disaccharides by *Chondroitinase ABC* and *Heparinase I-II-III*. The method omits proteolytic digestion, thereby limiting the derived depolymerized GAGs to the protein-free fraction – or free GAGs. Following depolymerization, disaccharides are labeled using 2-aminoacridone and injected into an UHPLC-MS/MS for separation and detection. The peaks of the 17 disaccharides are acquired at using multiple reaction monitoring analysis implemented in the mass spectrometry software (Waters® TargetLynx). The chromatographic conditions and MS configuration were set in accordance with the kit instruction for use.

Each sample was measured in singleton. The so-measured GAGome consisted of the absolute concentrations of 17 GAG disaccharides, corresponding to eight different sulfation patterns of chondroitin sulfate (CS) and heparan sulfate (HS), and one hyaluronic acid (HA) disaccharide. Specifically, we quantified eight CS disaccharides (0S CS, 2S CS, 6S CS, 4S CS, 2S6S CS, 2S4S CS, 4S6S CS, and TriS CS) and eight HS disaccharides (0S HS, 2S HS, 6S HS, NS HS, NS6S HS, NS2S HS, 2S6S HS, and TriS HS). We expanded the GAGome to include an additional 22 calculated features informative of GAG biology: a) the total CS and total HS concentration as the sum of the corresponding disaccharide concentrations, b) the CS charge [-] and HS charge [-] as the weighted sum of sulfated disaccharides, where the weight is the count of sulfo- groups in each disaccharide, c) two ratios (4S CS/0S CS and 6S CS/0S CS), and d) the relative concentration (or mass fraction, in µg/ µg %) of each of the 16 CS and HS disaccharides by normalizing each absolute concentration by the total CS and HS concentration, respectively. For each sample, the GAGome consisted of maximally 39 features.

We considered a GAGome feature detectable in plasma if the median concentration across all samples was above 0.1 µg/mL^25^. GAGome features that did not fulfill this criterion were excluded from downstream analyses.

### 2.4 cfDNA data

For 81 of the 113 samples, cfDNA concentration measurement was available. The cfDNA was extracted from 4 mL plasma using the QIAsymphony DSP Circulating DNA kit on the QIAsymphony SP system (Qiagen, Germany) according to the manufactureŕs instructions. The concentration was measured using dsDNA HS assay kit (Thermo Fisher) on Qubit 2.0 Fluorometer (Thermo Fisher). Out of the samples with available cfDNA concentration measurements, 74 samples also had NGS data available, which have been published previously^27^. The NGS panel used was AVENIO ctDNA Surveillance kit (Roche Diagnostics, Rotkreuz, Switzerland), which includes 197 cancer relevant genes.

### 2.5 Statistical analysis

Continuous data were presented using mean and median values while categorical data were presented using absolute and relative frequencies. Differences between LC patients and controls regarding general clinical characteristics were investigated using χ2-tests for categorical variables and Student’s t-tests for continuous data.

#### 2.5.1 GAGome analysis

We compared levels of each detectable GAGome feature in cancer versus control participants using Bayesian estimation with practical equivalence testing^28^. First, we fitted a Bayesian linear regression where each individual GAGome feature was standardized and modelled as a normally distributed response variable and the disease state (case vs control) was the only binary predictor. Second, we controlled the correlation between GAGome measurements in cancer vs controls for technical variation by fitting Bayesian linear models as above but including experimental batch (binary) and sample age (continuous, in months) as predictors. We used a t-distribution centered on 0 with 7 degrees of freedom, and scale = 2 for all the priors in all the Bayesian models. We fit the models using *rstanarm* package (ver 2.21.3) in R (ver. 4.2.1). The convergence and stability of the Bayesian sampling was assessed using R-hat, which should be below 1.01 ^29^ and Effective Sample Size (ESS), which should be greater than 1000^30^. The same convergence criteria were used for all Bayesian models.

A GAGome feature was considered credibly associated to case-control status if the 95% confidence interval (CI) of the difference in means did not include zero, and no more than 10% of the CI passed into the Region of Practical Equivalence (ROPE)^28^. The ROPE-interval was defined as [-0.1, 0.1] of the standardized mean, corresponding to a negligible effect size^31^. GAGome features that were credibly associated with case-control status after adjusting the linear regression model for experimental batch and sample age were further analyzed as predictors of cancer in the GAGome score development.

#### 2.5.2 GAGome score development

A Bayesian logistic model was fitted to predict case vs control using the absolute concentration of 0S CS (in µg/mL) and the fraction of 4S CS (in µg/µg %) as predictors. The predictors were batch-normalized before fitting the model. The Markov chain Monte Carlo (MCMC) sampling was performed with four chains of 10 000 iterations and a warmup of 5000. The output of the model is referred to as *plasma GAGome LC score,* or simply *GAGome score*.

Model metrics were investigated using a bootstrap analysis with 5000 bootstraps, under a constraint of minimum 95% specificity, which was deemed potentially clinically useful. The final model performance for detecting case vs control was evaluated by calculating sensitivity at 95% specificity, and an area under the curve (AUC) for the model.

#### 2.5.3 cfDNA score development

Bayesian logistic models (estimated using MCMC sampling) were fitted to predict case-control status using cfDNA concentration (logarithmic) in combination with the number of cfDNA variants. The output of the model is referred to as *cfDNA score*.

#### 2.5.4 Combined GAGome and cfDNA test pathway

A combined test was envisioned as a diagnostic pathway that sequentially uses the cfDNA score and the GAGome score to render a diagnostic decision, i.e. “combined test positive” vs “combined test negative”. To cumulatively retain the 95% specificity for the combined test, as was rationalized for the GAGome score, the GAGome specificity was kept at 95% while the specificity for the cfDNA score was set to 100%. Specifically, the GAGome score, when positive, could be used to reclassify cfDNA score false negative samples as “combined test positive”. The testing procedure was as following: 1) calculate the cfDNA score cut-off at which a minimum of 100% specificity was achieved, 2) calculate the GAGome score cut-off at which a minimum of 95% specificity was achieved, 3) for each sample, consider it positive for the cfDNA score, if the sample score was above the cut-off; analogously for the GAGome score, 4) for each sample, consider it positive for the combined test if at least one between the cfDNA and GAGome score were positive, 5) mark samples which were cfDNA score-negative but GAGome score-positive as “reclassified positive”.

## 3. Results

### 3.1 Cohort characteristics

The cohort comprised patients referred to the clinic under suspicion of LC (n=113). Of these, 85 cases were subsequently diagnosed with LC, which were further stratified into non-small cell LC (NSCLC, n=77) and small cell LC (SCLC, n=8). Patients diagnosed with a benign lung disease (n=28) served as controls (Table 1). Cases were slightly older than controls (mean 70.4 vs 67.0 years) and more evenly distributed among the sexes (52.9% females vs 60.7% females), neither characteristic showed a significant difference. A history of smoking was more prevalent among cases compared to controls (85.9% vs 60.8%, *p*=0.047). The predominant tumor stage was stage IV (61.2%), with a mean tumor size of 4.48 cm.

**Table 1.** Clinical characteristics of the cohort. Cases include patients diagnosed with lung cancer, irrespective of histology, and controls include patients diagnosed with different benign diseases of the lung.

| | Case<br>(n=85) | Control<br>(n=28) | Lung cancer vs<br>Controls, $p$-value | Overall<br>(N=113) |
| --- | --- | --- | --- | --- |
| **Age** |  |  |  |  |
| Mean (SD) | 70.4 (8.5) | 67.0 (14.4) | 0.246 | 69.6 (10.3) |
| **Gender** |  |  |  |  |
| Female | 45 (52.9%) | 17 (60.7%) | 0.473 | 63 (55.3%) |
| Male | 40 (47.1%) | 11 (39.3%) |  | 51 (44.7%) |
| **Smoking** |  |  |  |  |
| Smoker | 30 (35.3%) | 5 (17.9%) | 0.047 | 35 (31.0%) |
| Ex-Smoker | 43 (50.6%) | 12 (42.9%) |  | 55 (48.7%) |
| Never-Smoker | 12 (14.1%) | 9 (32.1%) |  | 21 (18.6%) |
| Missing | 0 (0%) | 2 (7.1%) |  | 2 (1.8%) |

|  | Case<br>(n=85) | Control<br>(n=28) | Lung cancer vs<br>Controls, *p*-value | Overall<br>(N=113) |
| --- | --- | --- | --- | --- |
| **Tumor size (cm)** |  |  |  |  |
| Mean (SD) | 4.5 (2.4) | NA (NA) |  | 4.5 (2.4) |
| Median [Min, Max] | 4.0 [0.9, 12.5] | NA [NA, NA] |  | 4.0 [0.9, 12.5] |
| Missing | 4 (4.7%) | 28 (100%) |  | 32 (28.3%) |
| **IASLC stage** |  |  |  |  |
| I | 9 (10.6%) | 0 (0%) |  | 9 (8.0%) |
| II | 6 (7.1%) | 0 (0%) |  | 6 (5.3%) |
| III | 18 (21.2%) | 0 (0%) |  | 18 (15.9%) |
| IV | 52 (61.2%) | 0 (0%) |  | 52 (46.0%) |

SD = Standard deviation, NA = Not Applicable and IASLC = International association for the study of lung cancer.

### 3.2 Correlation between plasma GAGomes and lung cancer diagnosis

The plasma GAGome, which comprised the structural characterization of 17 different GAG disaccharides, of all samples (n=113) was analyzed successfully. 4S CS and 0S CS were the only GAG disaccharides with a median detected concentration above 0.1 µg/mL. Therefore, we focused the subsequent analysis on these two independently measured features and the corresponding four derived features (Materials and methods, table S2, figure 1).

**Figure 1.**
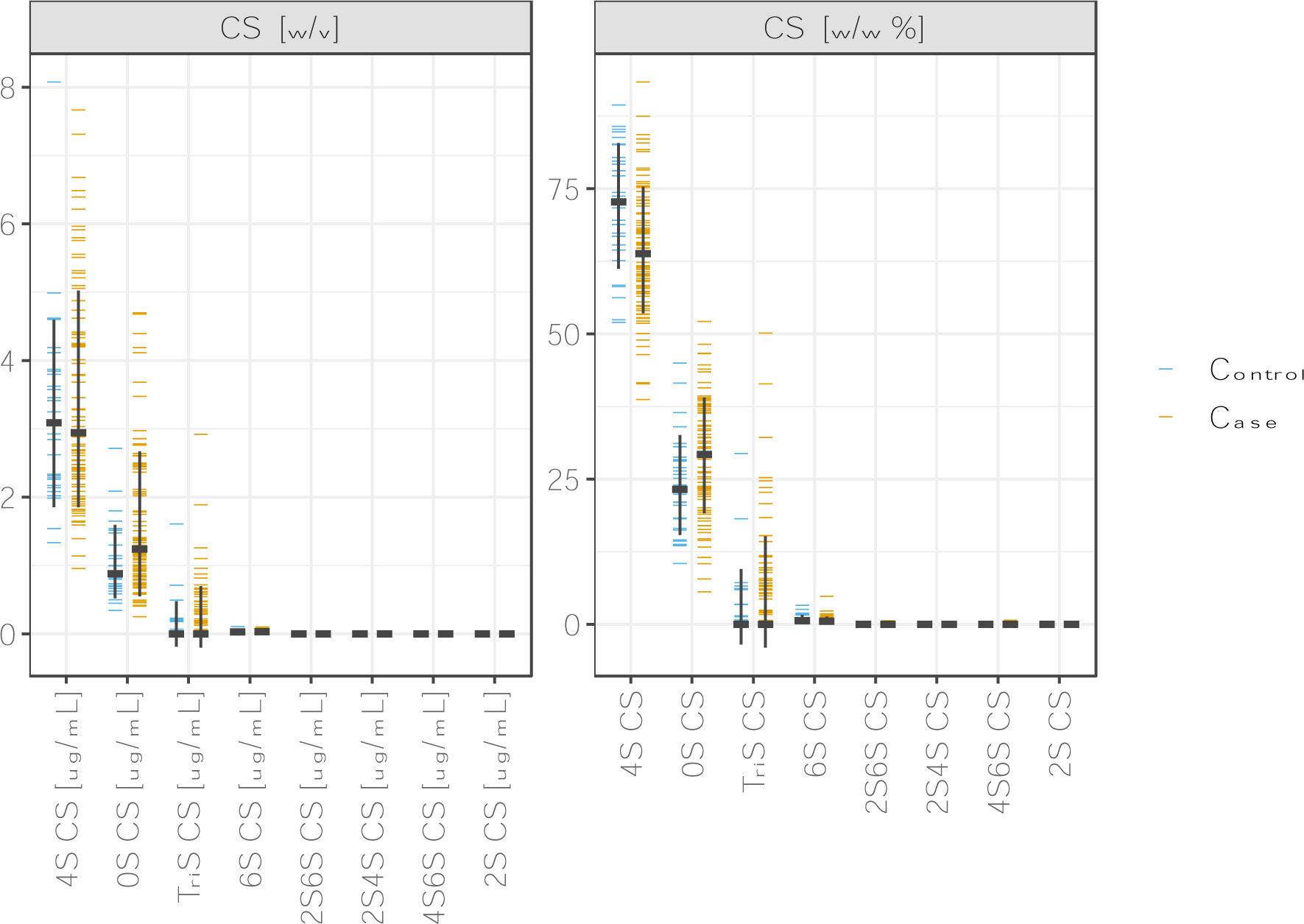
Plasma GAGome profiles. Concentrations (left) and fractions (right) of plasma GAGs in cases (n=85) and controls (n=28). CS: chondroitin sulfate.

Compared to controls, cases with cancer had a nominally higher median concentration of 0S CS, and a lower concentration of 4S CS (figure 1, table S2). Concomitantly, we found that the composition of CS in cases with cancer shifted towards a lower 4S CS fraction and higher 0S CS fraction (figure 1).

Using Bayesian linear regression and equivalence testing, we determined that two plasma GAGome features were credibly associated with LC after adjustment for batch effects and sample age (figure S1). Specifically, an increase in the plasma concentration of 0S CS and a reduction in the fraction of 4S CS were credibly correlated with LC after adjustment for confounders.

### 3.3 GAGome and cfDNA score

We next sought to develop a *plasma GAGome LC score* (or simply, GAGome score) for discriminating LC cases and controls. We fitted a Bayesian logistic model to predict cancer (figure S2, supplementary text), where the model’s output, referred to as the GAGome score, corresponded to the log-odds ratio of having LC.

The GAGome score had a 41.2% sensitivity (95% CI: 9.2-54.2%) to detect LC at a 96.4% specificity (95% CI: 95.2-100%) with an AUC of 0.67 (95% CI: 0.56-0.77, figure 2, table S3).

**Figure 2.**
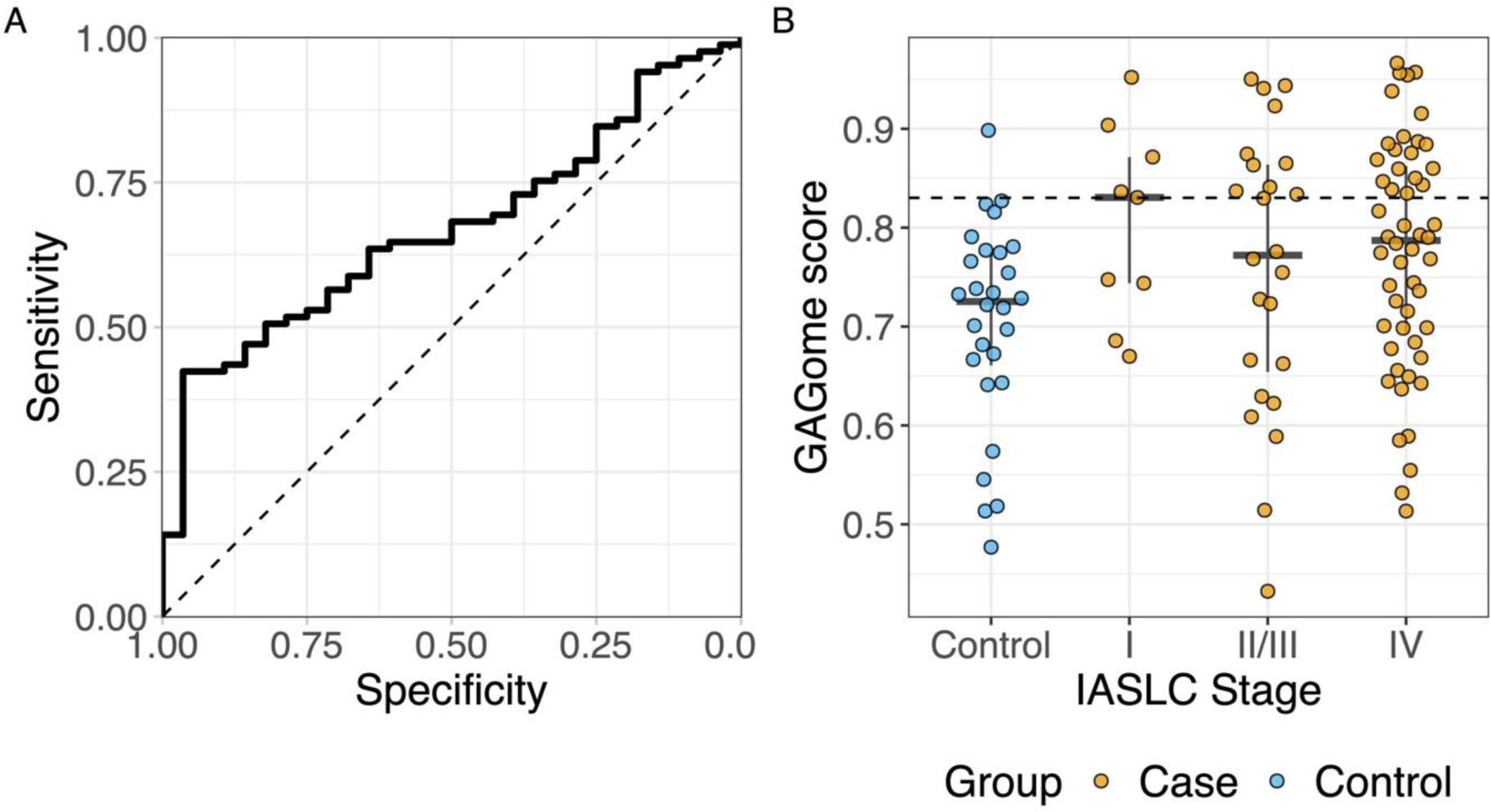
Performance of the GAGome score to discriminate cases and controls in A) the whole cohort and B) by IASLC stage. Controls are shown in blue (N_Control_ = 28) and cases in orange (N_Case_ = 85; N_StageI_ = 9, N_StageII/III_ = 24, N_StageIV_ = 58). IASLC: International Association for the Study of Lung Cancer.

Next, cfDNA data was analyzed. Neither cfDNA concentration nor number of cfDNA variants were correlated to the GAGome features included in the GAGome score (0S CS concentration and 4S CS fraction, figure S3). A *cfDNA score* was generated in the subset samples for which both cfDNA concentration and number of variants were available (n=74, figure 3A). Specifically, we fitted a Bayesian logistic model to predict case vs control using cfDNA concentration and the number of cfDNA variants as predictors. The model’s output, referred to as the cfDNA score, corresponded to the log-odds ratio of having LC. We treated samples where cfDNA concentration was available but insufficient for cfDNA variant analysis (n=7) as cfDNA score negative for evaluating the cfDNA score performance. The cfDNA score had an AUC of 0.80 (95% CI: 0.685-0.903) and a 42.6% sensitivity (95% CI: 31.7-60.6%) at 100% specificity (figure 3B).

**Figure 3.**
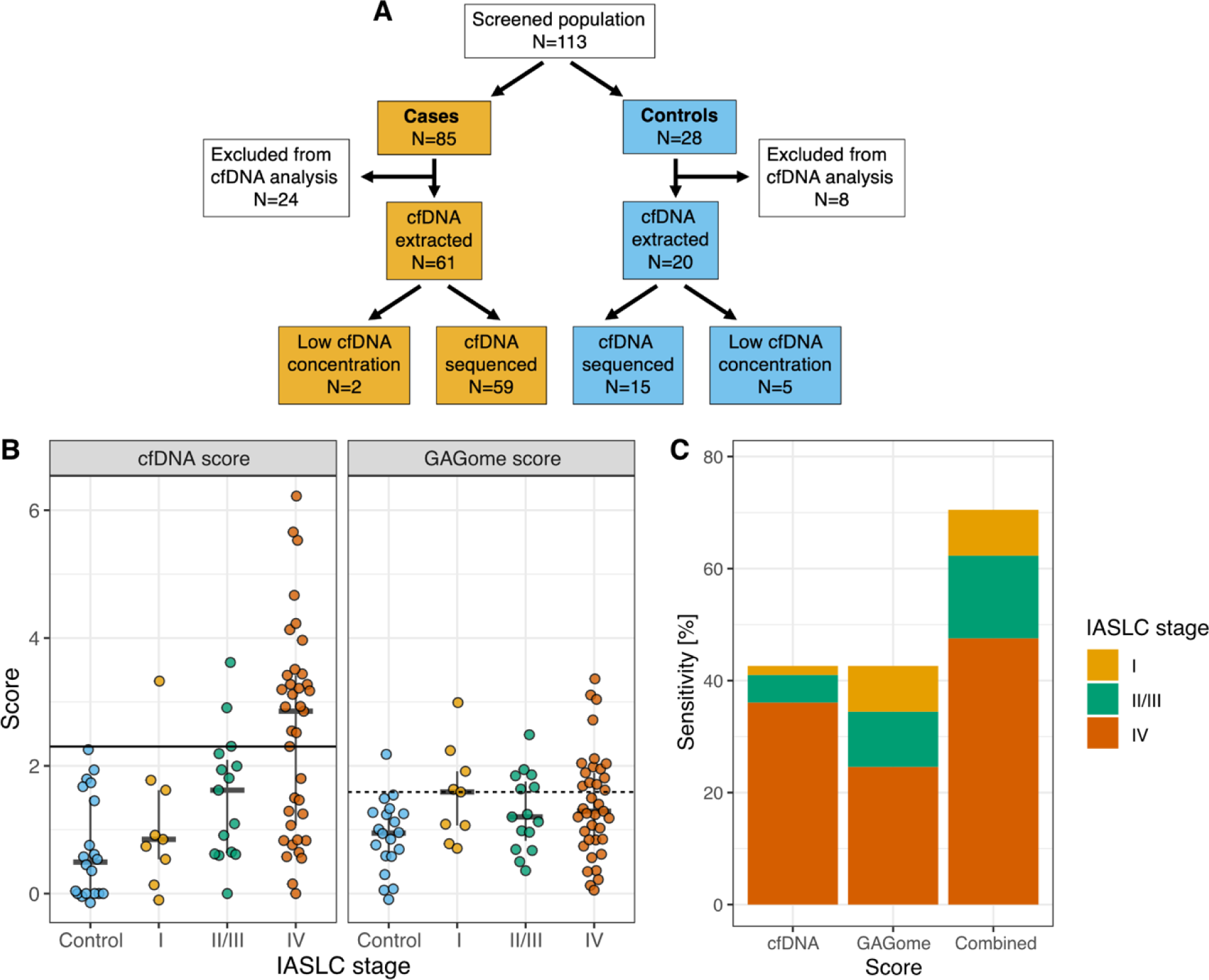
Performance of the combined test. A) Subject flow and inclusion for the cfDNA data. B) Performance and cutoffs for the cfDNA and GAGome score separately and C) sensitivity when combined (by IASCL stage). IASCL: International Association for the Study of Lung Cancer.

We envisioned a diagnostic pathway that relies on sequential cfDNA and GAGome measurements in plasma, specifically to use the GAGome score for potentially cfDNA score- false negatives. This combined test would be positive if either the cfDNA or GAGome scores were positive. We allowed for one false positive and set the threshold for cfDNA positivity at 100% specificity and GAGome score positivity at 95% specificity (figure 3). The sensitivity for the combined test increased to 70.5% (95% CI: 57.4 - 81.5%) at a specificity of 95% (95% CI: 75.1-100%, figure 3C). Notably, in this diagnostic pathway with overall 95% specificity (one false positive), the GAGome score contributed to a higher sensitivity for stage I (55.6% vs 11.1% for cfDNA alone, table S4) and stage II-III (40% vs 20% for cfDNA alone, table S4), and could correctly reclassify 17 out of 19 cases that were false negative when using the cfDNA score alone (table S5).

## 4. Discussion

In this study, we aimed to use GAGomes to discriminate between LC and benign lung diseases and to explore if the addition of cfDNA data could further increase the sensitivity of the developed score. We used the detected features, 0S CS and 4S CS, to build a GAGome score that reached a 41.2% sensitivity at 96.4% specificity. We next envisioned a test that combines the GAGome-score with cfDNA data to increase the diagnostic potential for LC. In the subset of patients with cfDNA data available the combined test could diagnose LC with 70.5% sensitivity at 95% sensitivity. To the best of our knowledge, this is the first study to develop a multiomics test combining GAGs and cfDNA for cancer.

Research on GAGs in LC has been limited, with most studies focusing on tissue-based analysis. An increase in total CS has been observed in tumor tissue compared to normal lung tissue^20,21,32^. However, the results regarding CS sulfation patterns have been inconsistent. Pál *et al*. and Balbisi *et al*. reported lower 0S CS and higher 4S CS in tumors, while Li *et al*. found the opposite for 4S CS. Discrepancies between tissue and plasma can be explained by the cell origin of the molecules analyzed. Mattox *et al*. investigated the contribution of different cells to cfDNA in plasma and found that, even in cancer patients, over 70% of cfDNA originated from leukocytes, and only 2.2% of the fragments in LC patients originated from the lung^33^. They concluded that these results reflect the systemic effect of the tumor on the body, suggesting that what is detected is not primarily the tumor itself, but the effect it causes. This concept can be applied to other circulating biomarkers, such as GAGs, indicating that plasma tests should be interpreted separately from tissue tests. Circulating GAGs in plasma has only been reported once previously^34^, where elevated 0S CS concentration and a lower 4S CS fraction was detected in LC cases relative to controls.

We found that the GAGome alone had a sensitivity of 40% for detecting LC which was comparable to 42.6% sensitivity of cfDNA. Given that GAGomes were uncorrelated with cfDNA measurements and that the GAGome score had similar sensitivity across all IASLC stages, we speculated that the two scores could be used in a combined multiomics test for LC. By combining scores derived from cfDNA and GAGs into a multiomic test we could show an increase in sensitivity to 70.5% for differentiating between LC and non-malignant lung diseases, highlighting the potential of a multiomics approach.

The effectiveness of using multiomics for cancer detection was notably proposed by Cohen *et al*. with CancerSEEK, which analyzed mutations in cfDNA and protein levels^3^. They reported an overall sensitivity of 70% across all tumor types and about 59% for LC specifically. Since then, several other studies have explored the potential of multiomics as a diagnostic tool.

For instance, Wang *et al*.^4^ tested a combination of cfDNA variants, proteins and fragmentomics on a cohort of colorectal, esophageal, gastric, liver, lung, and ovarian cancers ^35^. At 98% specificity, cfDNA variants alone showed a sensitivity of 46%. This increased to 60% with the addition of proteins, and further to 66% with fragmentomics. However, for LC alone, the sensitivity of the combined model only reached 38.5%. Chen *et al*. combined a mutational score, methylation, and serum CEA levels for distinguishing between LC and benign lung nodules, achieving a 76.9% sensitivity at a modest specificity of 58.3%. At the 95% specificity level used in our study, their sensitivity dropped to around 30%^6^. D’Ambrosi *et al*. reported a promising 68% sensitivity at 95% specificity for diagnosing LC using platelet-derived circRNA and mRNA^36^. Notably, their control group primarily consisted of asymptomatic individuals, similar to Wang *et al*. and CancerSEEK. Our study differed in design by using patients referred to the clinic with lung-related symptoms qualifying for LC evaluation, rather than selecting a specific control group. This enhances the clinical relevance of our findings, as it better reflects real-world diagnostic challenges. Although our control cohort was more suitable compared to several other multiomics studies, it is important to note that most of these studies had a skew towards lower stages of LC, which may have negatively impacted their reported sensitivity.

The only detectable plasma GAGome features in this cohort were 0S CS and 4S CS. This is consistent with previous findings using this kit, both in healthy individuals and cancer^34,37^. Other studies have detected additional GAGs, such as HA in plasma^38^ . However, the methods used in those studies included proteolytic digestion, while the method used here is degradation-free, detecting only GAGs that circulate freely in the analyzed liquid. Saito *et al.* tested the affinity of plasmatic proteins to different GAGs and could show that approximately 7.5% of the proteins are bound to HS and dermatan sulfate, while only 0.25% are bound to CS^39^ . This suggests that CS has a higher probability of circulating freely in plasma, supporting our findings.

A limitation of our study includes the relatively small sample size and lack of external cohorts for validation. A methodological limitation is that blood was collected in tubes optimized for stabilization of cells and extracellular RNA, and not immediately processed, as the stabilization reagents are effective for several days. However, GAGs are primarily degraded by highly substrate-specific enzymes in the lysosome^40^. Since the tubes’ main function is to stabilize cells, lysosomal activity in the plasma fraction of the sample is unlikely. While an unspecific degradation in plasma cannot be ruled out, the cancer-specific alterations to GAGomes were robust to adjustment for sample age.

In conclusion, we have shown that free CS can be detected in the plasma of LC patients. From this, we developed highly specific and sensitive multiomics score by combining this data with cfDNA data from NGS analysis, effectively differentiating between LC and benign lung diseases. We envision this score as a natural companion diagnostic to radiography as a broad NGS is highly relevant for LC due to the increasing number of targeted therapies available for this indication. Adding a GAGome analysis would be comparatively inexpensive, easily performed on a small aliquot of the same blood sample used for NGS analysis and could specifically increase sensitivity for detecting lower stages of cancer without affecting the false positive rate.

## Data availability

Data is available from the authors upon reasonable request.

