## Supplementary for "Plasma glycosaminoglycans and cell-free DNA to discriminate benign and malignant lung diseases"

Family name followed by given name: Qvick Alvida[1], Bratulic Sinisa[2], Carlsson Jessica[3],

Stenmark Bianca[1], Karlsson Christina[4], Nielsen Jens[2,5], Gatto Francesco[2,6], Helenius Gisela[1]

### Supplementary text

#### GAGome score

Following the Sequential Effect eXistence and sIgnificance Testing (SEXIT) framework, we reported the median of the posterior distribution and its 95% CI (Highest Density Interval), along the probability of direction (pd), the probability of significance, and the probability of being large. The thresholds beyond which the effect is considered as significant (i.e., non-negligible) and large are |0.09| and |0.54|. Convergence and stability of the Bayesian sampling has been assessed using Rhat, which should be below 1.01 (Vehtari *et al.*, 2019), and Effective Sample Size (ESS), which should be greater than 1000 (Burkner, 2017).

We fitted a Bayesian logistic model to predict cancer with the formula: cancer ~X0s_CS_plasma_conc + X4s_CS_plasma (supplemental 1). The model's explanatory power was weak (R2 = 0.08, 95% CI [7.65e-03, 0.17]). The model's intercept was at 1.26 (95% CI [0.81, 1.77]). Within the model, the effect of X0s CS plasma concentration ($\beta_1$= 0.51, 95% CI [-0.06, 1.18]) had a 95.87% probability of being positive (> 0), 92.10% of being significant (> 0.09), and 45.28% of being large (> 0.54). The estimation successfully converged (Rhat = 1.000) and the indices were reliable (ESS = 15210). The effect of X4s CS plasma ($\beta_2$= -0.43, 95% CI [-0.94, 0.03]) had a 96.97% probability of being negative (< 0), 92.47% of being significant (<-0.09), and 32.70% of being large (< -0.54). The estimation successfully converged (Rhat = 1.000) and the indices were reliable (ESS = 17116).

### Supplementary Figures

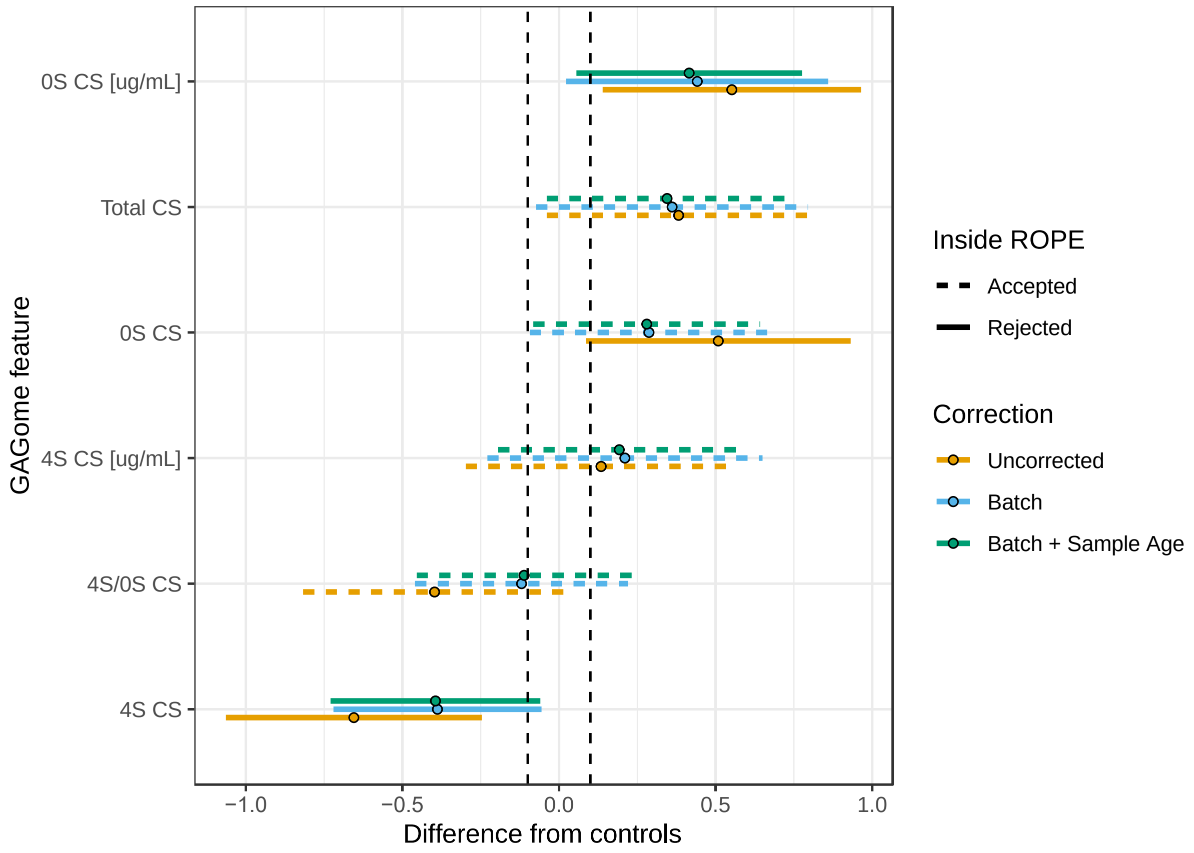


**Figure S1.** Posterior distributions of differences in GAGome features between cases and controls in Bayesian region of practical equivalence (ROPE) testing for linear regression models with and without correction for technical effects. Dots show medians and lines show 95% credible intervals (CI). Greyed area represents ROPE corresponding to the [-0.1,0.1] interval on the standardized mean. Practical equivalence to zero is rejected (solid lines) if less than 5% of the distribution's CI falls within the ROPE interval and accepted otherwise (dashed line).

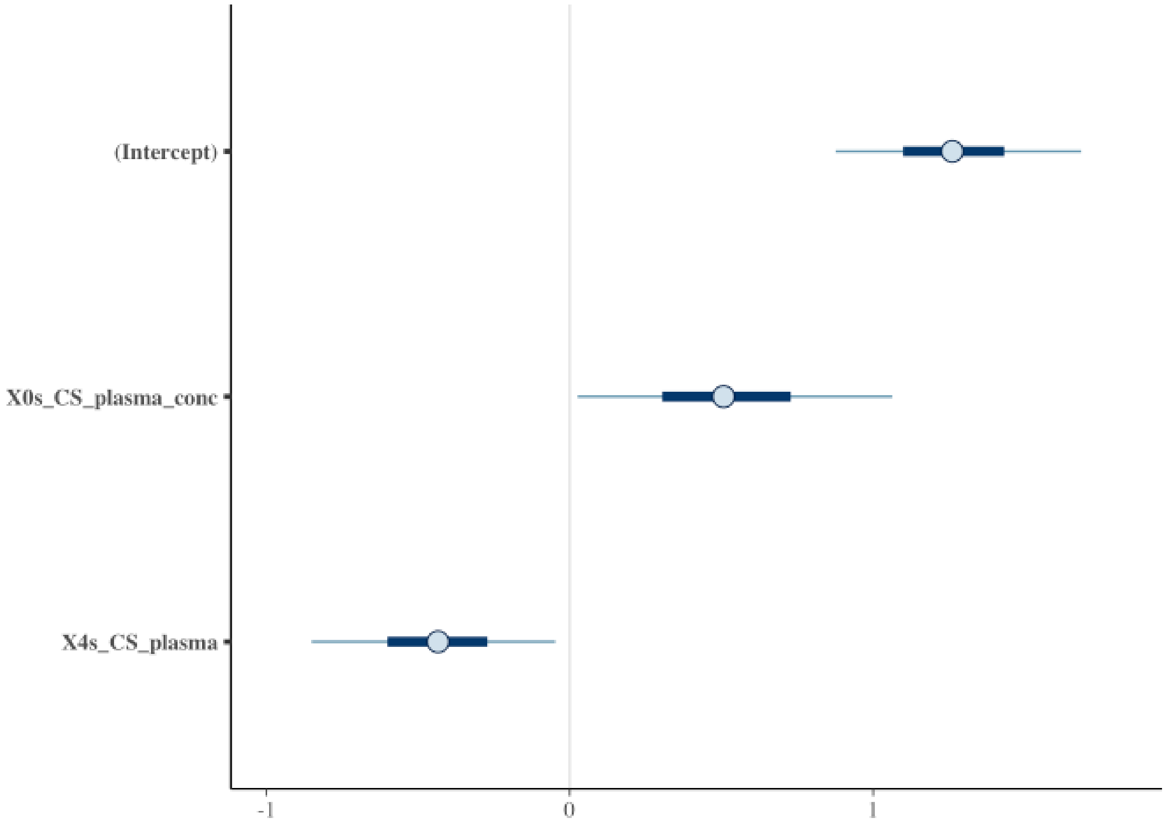


**Figure S2.** GAGome score model summary with Bayesian logistic regression coefficients. The plot shows the predictors' median (circle) as well as 50% (thick horizontal line) and 95% (thin horizontal line) CI.

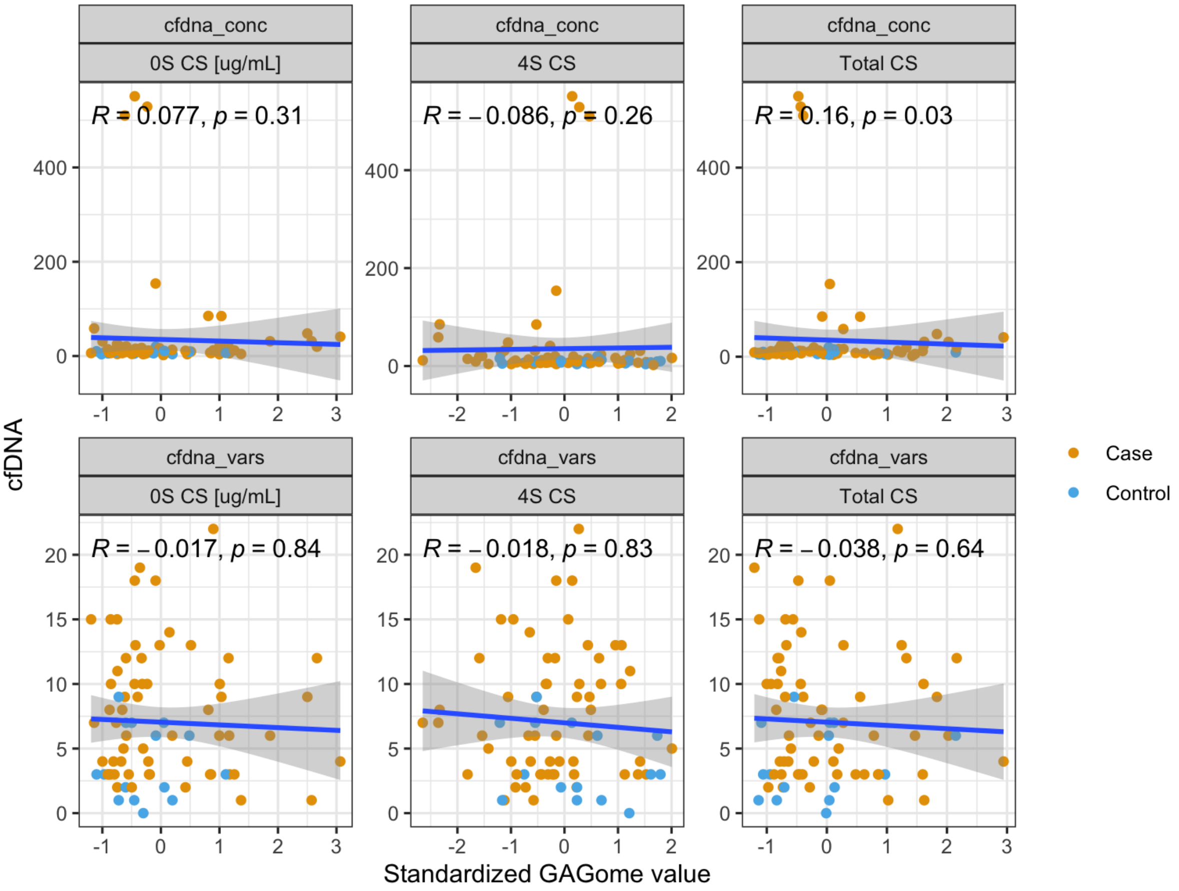


**Figure S3.** Correlations between cfDNA concentrations (top)/cfDNA variant count (bottom) and GAGome score components expressed as Kendall correlation coefficients and corresponding *p*-values.

### Supplementary Tables

**Table S1.** STARD checklist

| Section & Topic | No | Item | Reported on page # |
|---|---|---|---|
| **TITLE OR ABSTRACT** | | | |
| | **1** | Identification as a study of diagnostic accuracy using at least one measure of accuracy (such as sensitivity, specificity, predictive values, or AUC) | 2 |
| **ABSTRACT** | | | |
| | **2** | Structured summary of study design, methods, results, and conclusions (for specific guidance, see STARD for Abstracts) | 2 |
| **INTRODUCTION** | | | |
| | **3** | Scientific and clinical background, including the intended use and clinical role of the index test | 3-4 |
| | **4** | Study objectives and hypotheses | 4 |
| **METHODS** | | | |
| *Study design* | **5** | Whether data collection was planned before the index test and reference standard were performed (prospective study) or after (retrospective study) | 5 |
| *Participants* | **6** | Eligibility criteria | 4-5 |
| | **7** | On what basis potentially eligible participants were identified (such as symptoms, results from previous tests, inclusion in registry) | 4-5 |
| | **8** | Where and when potentially eligible participants were identified (setting, location and dates) | 4 |
| | **9** | Whether participants formed a consecutive, random or convenience series | 4 |
| *Test methods* | **10a** | Index test, in sufficient detail to allow replication | 5-9 |
| | **10b** | Reference standard, in sufficient detail to allow replication | 4-5 |
| | **11** | Rationale for choosing the reference standard (if alternatives exist) | 4 |
| | **12a** | Definition of and rationale for test positivity cut-offs or result categories of the index test, distinguishing pre-specified from exploratory | 6, 8-9 |
| | **12b** | Definition of and rationale for test positivity cut-offs or result categories of the reference standard, distinguishing pre-specified from exploratory | 4 |
| | **13a** | Whether clinical information and reference standard results were available to the performers/readers of the index test | 5 |
| | **13b** | Whether clinical information and index test results were available to the assessors of the reference standard | 5 |
| *Analysis* | **14** | Methods for estimating or comparing measures of diagnostic accuracy | 7-9 |
| | **15** | How indeterminate index test or reference standard results were handled | 4, 6 |
| | **16** | How missing data on the index test and reference standard were handled | 4, 6 |
| | **17** | Any analyses of variability in diagnostic accuracy, distinguishing pre-specified from exploratory | |
| | **18** | Intended sample size and how it was determined | 4 |
| **RESULTS** | | | |
| *Participants* | **19** | Flow of participants, using a diagram | 14 |
| | **20** | Baseline demographic and clinical characteristics of participants | 9-10 |
| | **21a** | Distribution of severity of disease in those with the target condition | 10 |
| | **21b** | Distribution of alternative diagnoses in those without the target condition | 4 |
| | **22** | Time interval and any clinical interventions between index test and reference standard | NA |
| *Test results* | **23** | Cross tabulation of the index test results (or their distribution) by the results of the reference standard | Supplementary |
| | **24** | Estimates of diagnostic accuracy and their precision (such as 95% confidence intervals) | 12, 13 |
| | **25** | Any adverse events from performing the index test or the reference standard | NA |
| **DISCUSSION** | | | |

| | | | |
|---|---|---|---|
| | **26** | Study limitations, including sources of potential bias, statistical uncertainty, and generalisability | 17 |
| | **27** | Implications for practice, including the intended use and clinical role of the index test | 17 |
| **OTHER INFORMATION** | | | |
| | **28** | Registration number and name of registry | NA |
| | **29** | Where the full study protocol can be accessed | Full details presented in manuscript |
| | **30** | Sources of funding and other support; role of funders | 18 |

**Table S2.** Detectable plasma GAGome features in the included population.

| Concentration (µg/mL) | Case (n=85) | Control (n=28) | Overall (N=113) |
|---|---|---|---|
| **Total CS** | | | |
| Mean (SD) | 5.34 (2.36) | 4.47 (1.72) | 5.12 (2.24) |
| Median [Min, Max] | 4.53 [1.84, 12.42] | 4.48 [2.55, 10.19] | 4.53 [1.84, 12,42] |
| **0S CS** | | | |
| Mean (SD) | 1.61 (1.06) | 1.06 (0.54) | 1.47 (0.99) |
| Median [Min, Max] | 1.24 [0.25, 4.70] | 0.88 [0.34, 2.72] | 1.13 [0.25, 4.70] |
| **4S CS** | | | |
| Mean (SD) | 3.44 (1.59) | 3.23 (1.37) | 3.38 (1.53) |
| Median [Min, Max] | 2.94 [0.96, 7.67] | 3.09 [1.34, 8.08] | 2.94 [0.96, 8.08] |

**Table S3.** GAGome score sensitivity per stage at 95% specificity in the total population.

| IASLC Stage | Total | True positive | False negative | Sensitivity |
|---|---|---|---|---|
| I | 9 | 5 | 4 | 55.6% |
| II | 6 | 4 | 2 | 66.7% |
| III | 18 | 6 | 12 | 33.3% |
| IV | 52 | 20 | 32 | 38.5% |

IASLC = International association for the study of lung cancer.

**Table S4.** cfDNA, GAGome and combined test performance across stages in the subset of cases with available cfDNA. Samples (n=2) with cfDNA concentration available but insufficient for cfDNA variant analysis in the cfDNA test were assigned as negative, corresponding to one sample in stage II/III and one in stage IV group.

| | **cfDNA test (100% specificity)** | | | **GAGome test (95% specificity)** | | | **Combined test (95% specificity)** | | |
|---|---|---|---|---|---|---|---|---|---|
| | **Stage I (N=9)** | **Stage II/III (N=15)** | **Stage IV (N=37)** | **Stage I (N=9)** | **Stage II/III (N=15)** | **Stage IV (N=37)** | **Stage I (N=9)** | **Stage II/III (N=15)** | **Stage IV (N=37)** |
| NEGATIVE | 8 (88.9%) | 12 (80.0%) | 15 (40.5%) | 4 (44.4%) | 9 (60.0%) | 22 (59.5%) | 4 (44.4%) | 6 (40.0%) | 8 (21.6%) |
| POSITIVE | 1 (11.1%) | 3 (20.0%) | 22 (59.5%) | 5 (55.6%) | 6 (40.0%) | 15 (40.5%) | 5 (55.6%) | 9 (60.0%) | 29 (78.4%) |

**Table S5.** The multiomics diagnostic pathway results in the subset of cases with available cfDNA. The GAGome test, when positive, is used to reclassify subjects who were negative or inconclusive on the cfDNA test. Samples with cfDNA concentration available but insufficient for cfDNA variant analysis are listed as Inconclusive for the cfDNA test.

| **Arm** | **cfDNA test** | **GAGome test** | **Combined test +** | **Combined test -** | **Reclassified as +** |
|---|---|---|---|---|---|
| Case | Inconclusive | Negative | 2 | 0 | FALSE |
| Case | Negative | Negative | 16 | 0 | FALSE |
| Case | Negative | Positive | 0 | 17 | TRUE |
| Case | Positive | Negative | 0 | 17 | FALSE |
| Case | Positive | Positive | 0 | 9 | FALSE |
| Control | Inconclusive | Negative | 5 | 0 | FALSE |
| Control | Negative | Negative | 14 | 0 | FALSE |
| Control | Negative | Positive | 0 | 1 | TRUE |
